# EPIDEMIOLOGY OF MAXILLOFACIAL INJURIES IN “HERATSI” NO 1 UNIVERSITY HOSPITAL IN YEREVAN, ARMENIA: A RETROSPECTIVE STUDY

**DOI:** 10.1101/2021.06.02.21258208

**Authors:** Lusine V. Aleksanyan, Anna Yu. Poghosyan, Martin S. Misakyan, Armen M. Minasyan, Aren Yu. Bablumyan, Artashes E. Tadevosyan, Armen A. Muradyan

**Affiliations:** Department of ENT and Maxillofacial Surgery, Yerevan State Medical University, “Heratsi” No 1 Hospital, 60 Abovyan Str., Yerevan, Armenia, 0025; Administrative Department, Yerevan State Medical University, 2 Koryun Str., Yerevan, Armenia, 0025; Department of Public Health and Healthcare, Yerevan State Medical University, 2 Koryun Str., Yerevan, Armenia, 0025

**Keywords:** Epidemiology, Maxillofacial fracture, Mandible, Etiology, Interpersonal violence

## Abstract

**Objective:** The aim of this study was to perform a retrospective analysis of the prevalence, etiologies, and types of maxillofacial injuries (MFI) and sites of maxillofacial fractures (MFF) and their management in the Department of ENT and Maxillofacial Surgery of <<Heratsi>> No. 1 University Hospital in Yerevan, Armenia.

**Material and methods:** A retrospective cross-sectional study was conducted. Data including age, sex, date of referral, mode of injury, etiology, radiology records and treatment methods were extracted. Study outcomes were measured using percentages, means, standard deviations and tests of proportions. P <.05 was considered significant.

**Results:** A total of 204 patients had a mean age of 36.26 ±1.08 years (156 males and 48 females), and a total of 259 MFIs were recorded between 2017 and 2020. Interpersonal violence was found to be the most common etiology of MFFs in this study (42.1%), followed by road traffic accidents (RTAs) (27.9%) and falls (18.6%). The nasal bone was the most common injury site (47.5%), followed by the mandible (31.4%) and zygomatic complex (11.7%). The most common fracture site was the mandibular angle (37.9%), followed by the symphysis/parasymphysis (28.1%) and body (12.6%). The majority of MFFs were treated by open reduction and internal fixation.

**Conclusion:** Interpersonal violence, followed by RTAs and falls, was the most common cause of MFIs. The nasal bone was the most common injury site, followed by the mandible and zygomatic complex. Social education with the objective of reducing aggression and interpersonal conflict should be improved, and appropriate RTA prevention strategies should be strengthened and implemented.

**KEY MESSAGES:** *What is already known on this subject:*

- Traumatic injuries continue to be important causes of morbidity and mortality in both developed and developing countries.
- The incidence rates, etiologies, types, and injuries associated with MFFs vary among different countries.
- RTAs are the most frequent cause of MFIs in developing countries, and in contrast, the most frequent cause of MFFs in developed countries is IV or assault

*What this study adds:*

- Interpersonal violence was found to be the most common etiology of MFF in this study, followed by RTAs and falls.
- Patients aged 21-30 years were likely to have sustained nasal bone fractures and mandible fractures in equal proportions.
- The nasal bones were found to be the most common injury site, followed by the mandible and zygomatic complex.

## INTRODUCTION

Traumatic injuries continue to be important causes of morbidity and mortality in both developed and developing regions.[1-4] The epidemiology of facial injuries varies among different countries and geographic zones. Population concentration, lifestyle, cultural background, and socioeconomic status can affect the prevalence of maxillofacial injuries (MFIs).[4-7] In addition to population and societal changes, the incidence rates and patterns of maxillofacial fractures (MFFs) may also vary among time periods due to legislative changes such as the introduction of compulsory safety belt legislation, helmet use, and speed limit enforcement.[6-10] Traumatic injuries represent a significant and growing disease burden in the developing world and are now one of the leading causes of death in economically active adults in many low- and middle-income countries.[4, 9, 11] According to the World Health Organization (WHO), middle-income countries have higher injury and death rates than low- and high-income countries.[3, 12] In addition an increasing total proportion of injuries in developing countries, among the total number of injuries to the maxillofacial region, the percentage of combined injuries is increasing, which indicates serious suffering among patient and prolonged hospitalization and rehabilitation.[2, 13-15]

Maxillofacial fractures can be considered consequential injuries, as they may result in mortality, severe morbidity, facial disfigurement, and functional limitations.[2] Knowledge about the epidemiology of MFF can help practitioners make appropriate clinical decisions and guide professionals and policy makers concerned with developing suitable injury prevention strategies.

The aim of this study was to perform a retrospective analysis of the prevalence, etiologies, and types of maxillofacial injuries and sites of maxillofacial fractures and their management in the Department of ENT and Maxillofacial Surgery of <<Heratsi>> No. 1 University Hospital in Yerevan, Armenia.

## MATERIAL AND METHODS

This research was conducted in accordance with relevant ethical standards, and the study protocol was approved by the Yerevan State Medical University Ethics Committee [IRB №5-3/2021]

### Patient and Public Involvement

Patients and the public were not involved in any way

A retrospective cross-sectional study was conducted. The medical records of hospitalized patients with MFIs admitted to the Department of ENT and Maxillofacial Surgery at <<Heratsi>> No. 1 University Hospital in Yerevan, Armenia, between January 2017 and December 2020 were retrieved and analyzed to obtain prevalence, etiology, injury pattern and treatment modality data.

The exclusion criteria were as follows: 1) outpatients offered immediate treatment without hospitalization; 2) patients with only soft tissue injuries who were treated in the emergency room without hospitalization; or 3) military patients wounded during the war from October-November 2020.

After excluding such patients, the records of 204 patients aged between 12 and 90 years were retrospectively analyzed. The sample size calculation n = Z^2^pq/ Δ^2^ was performed for a one group proportion, where p=0.5, Δ=0.07, n=196.

Data on age, sex, date of referral, mode of injury, etiology, radiographic findings with radiology records and treatment methods were extracted. Injury etiology was classified into four main categories: (1) RTAs involving automobiles, motorcycles and bicycles, including drivers, pillion riders, passengers, and pedestrians; (2) falls from heights, household falls, and falls due to systemic illness such as epilepsy or while playing; (3) assaults or interpersonal violence; and (4) sport-related and other injuries.

The type of MFF was classified according to the following maxillofacial anatomical sites: nasal (N), Le-Fort (LeF), zygomatic complex (ZC), orbital floor (OF) and mandibular (M) (subclassified into symphysis/parasymphysis, body, angle, ramus, condylar and coronoid process) fractures.

MFIs were treated with the following methods: 1) closed reduction (CR); 2) open surgical treatment or open reduction and internal fixation (ORIF), conservative treatment (CT) and wound debridement (WD).

Data collection tools consisted of observation and census sampling of medical records and documents.

### Ethical considerations

Ethical considerations were taken into account throughout the study, and the patients’ names and medical information were kept completely confidential. The subjects’ medical history was used solely for the purposes of the current study.

### Statistical analysis

Statistical analysis was performed using SPSS version 16.0 (SPSS Inc., Chicago, IL, USA). Study outcomes were measured using percentages, means, standard deviations and tests of proportions. The prevalence rates of injuries in particular age, sex, etiology, and fracture type groups were analyzed. Non parametric statistic test (Pearson’ χ^2^) was used describing nominal data. Only for description of patients age mean and SD was applied. Patient- and injury-related variables, including age, sex, anatomic location of the fracture, and etiology, were analyzed with χ^2^ tests or tables larger than 2×2, a post hoc test with Bonferroni correction was used.

## RESULTS

From 2017-2019, MFIs increased annually (Figure 1). In 2020, the total number of injuries decreased because of some restrictions and lockdowns in Armenia due to the coronavirus disease 2019 (COVID-19) pandemic. Despite the absence of strict patterns, it was observed that the highest rate of fractures occurred from July to October (Figure 1).

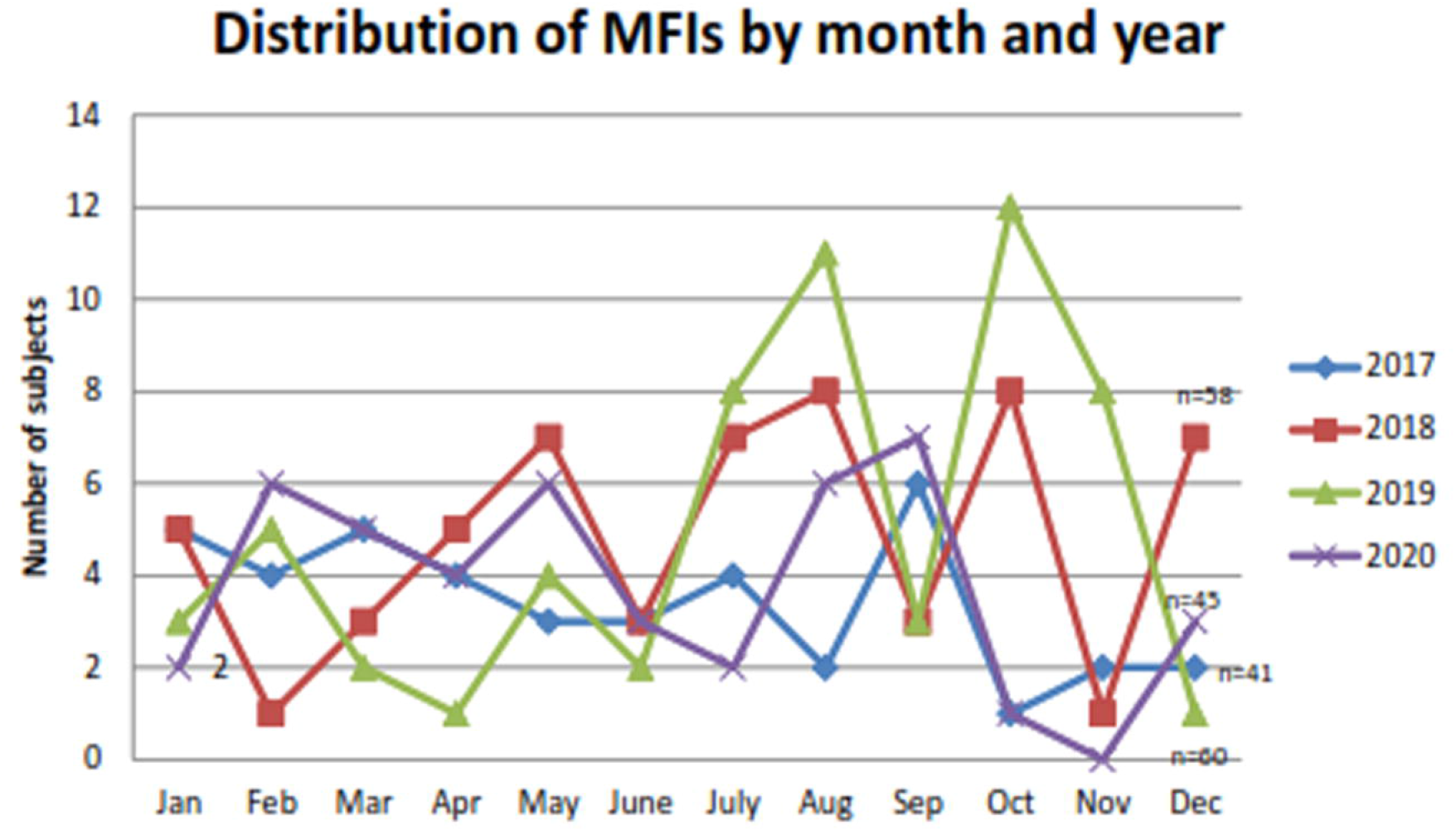

A total of 204 patients with 259 MFIs presented to the ENT and Maxillofacial Surgery Department between 2017 and 2020.

Patients with MFFs accounted for 190 of the 204 patients (93.14%), with a total of 242 fractures.

The mean age and standard deviation of the patients with MFIs was 36.26 ±1.08 years, with a minimum age of 12 years and maximum age of 90 years. Adults aged between 21 and 40 years had the highest rate. In this study, 76.5% (156/204) of the subjects were male, and 23.5% (48/204) were female, with a male to female ratio of 3:1. The test of proportion of males and females showed that there was a significantly higher proportion of males with maxillofacial trauma (*P*=0.0009, n=204, χ^2^ test).

As presented in Figure 2, males in the 21-30 years age group had the highest prevalence (33.4%; n=68). The highest prevalence in females occurred in the age groups over 61 years (6.4%; n=13).

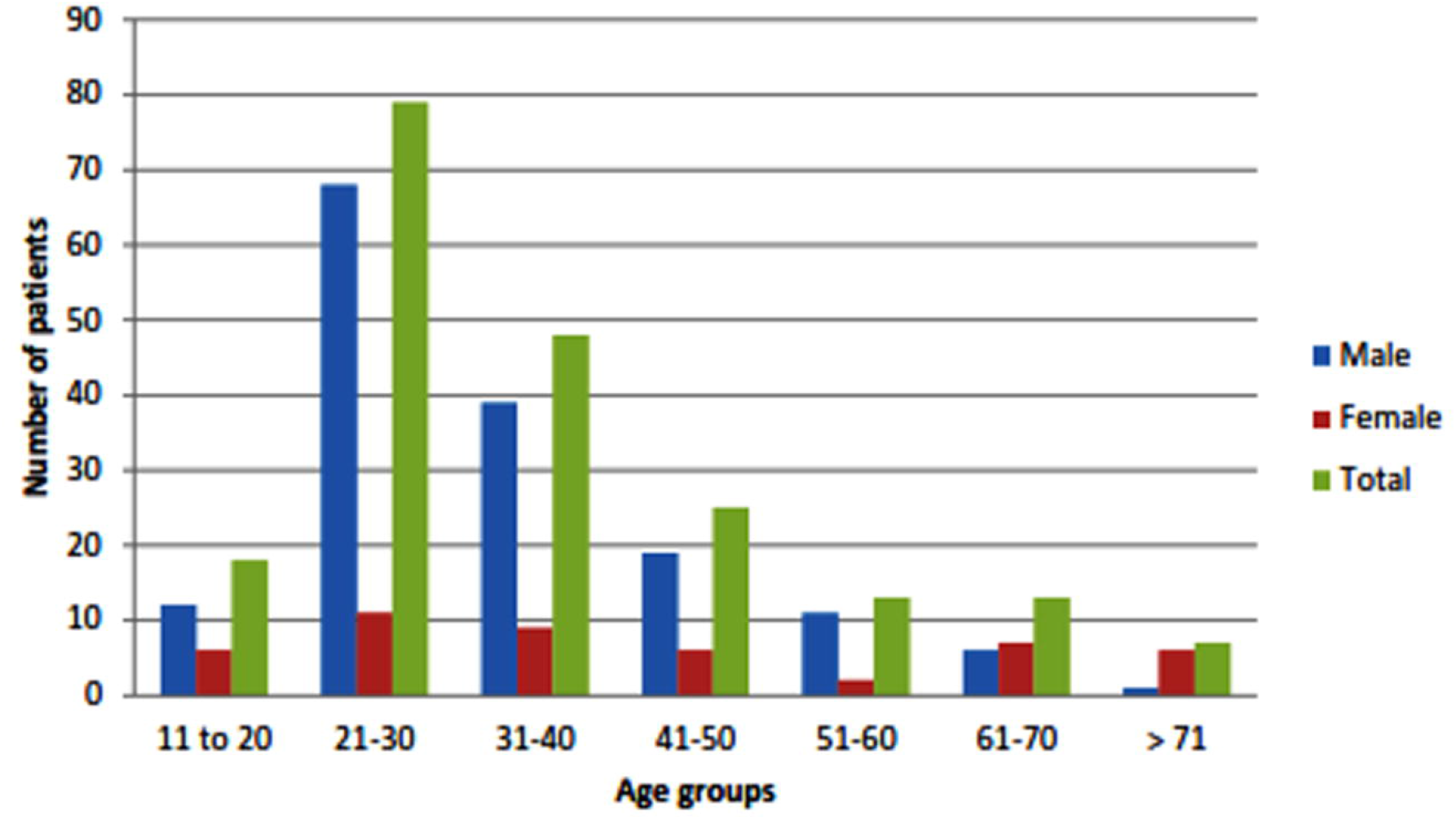

The most common cause of MFI was interpersonal violence (IV), accounting for 42.1% of all injuries (86/204), and male predominance was observed ((94.1% vs 1.1%; p= 0.0006, n=204, χ^2^ test). RTAs accounted for 27.9% (57/204) of all injuries (Figure 3). Car accidents and pedestrian accidents were the main causes of RTA injuries, and only one motorcycle accident was reported. Trauma due to falls accounted for 18.6% of the injuries (38/204), mostly involving females (52.6%; 20/38), elderly people who fell due to systemic illness and men who fell from heights. Domestic injuries accounted for 5.4% of all injuries (11/204). Six sports-related injuries, five industrial injuries and one suicide-related injury were reported.

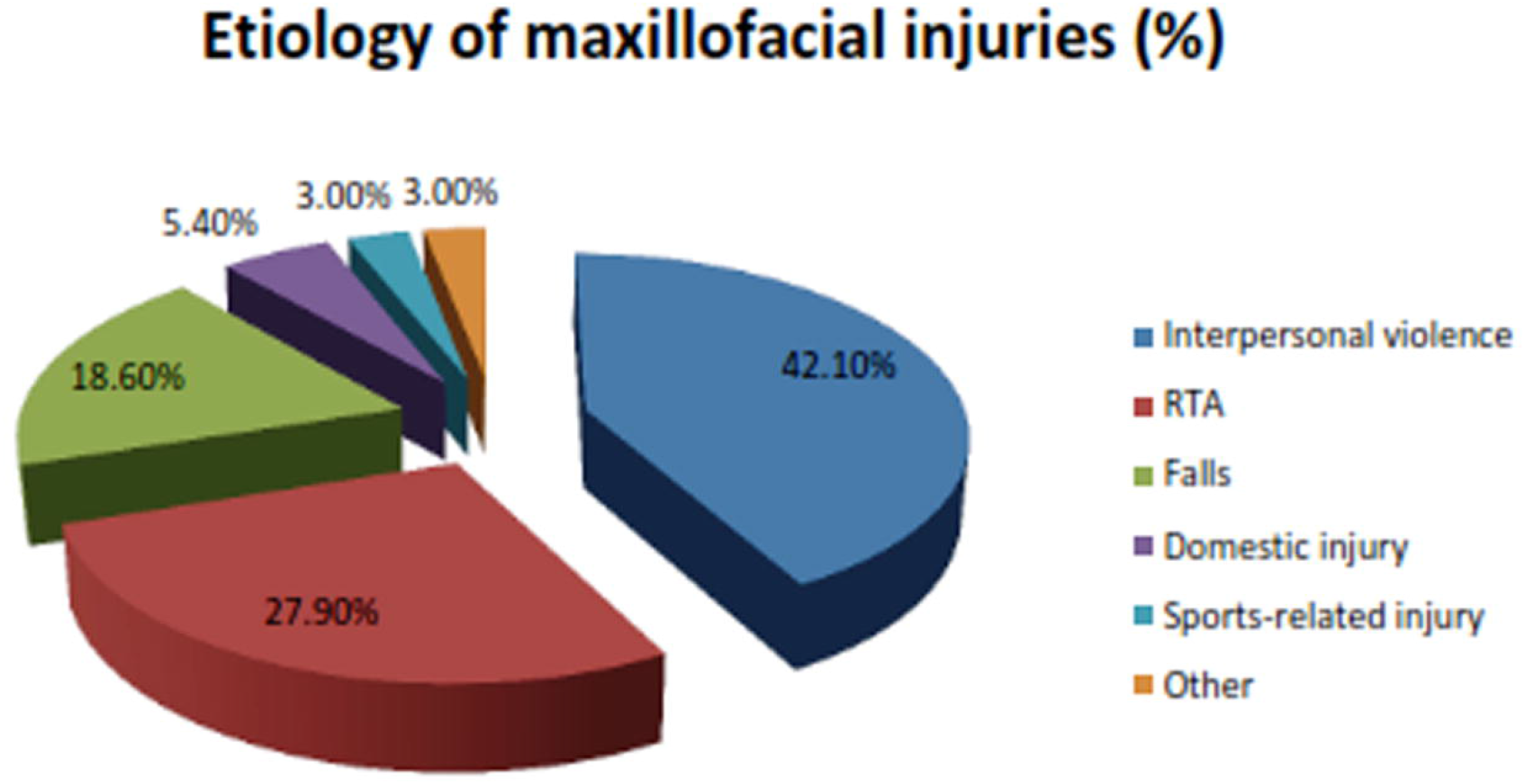

Data analysis showed that the largest percentage of fractures occurred in the nasal bones, accounting for 47.5% of all MFIs (n=204), of which 82 were isolated fractures of the nose and 15 were combined with other maxillofacial fractures (Figure 4).

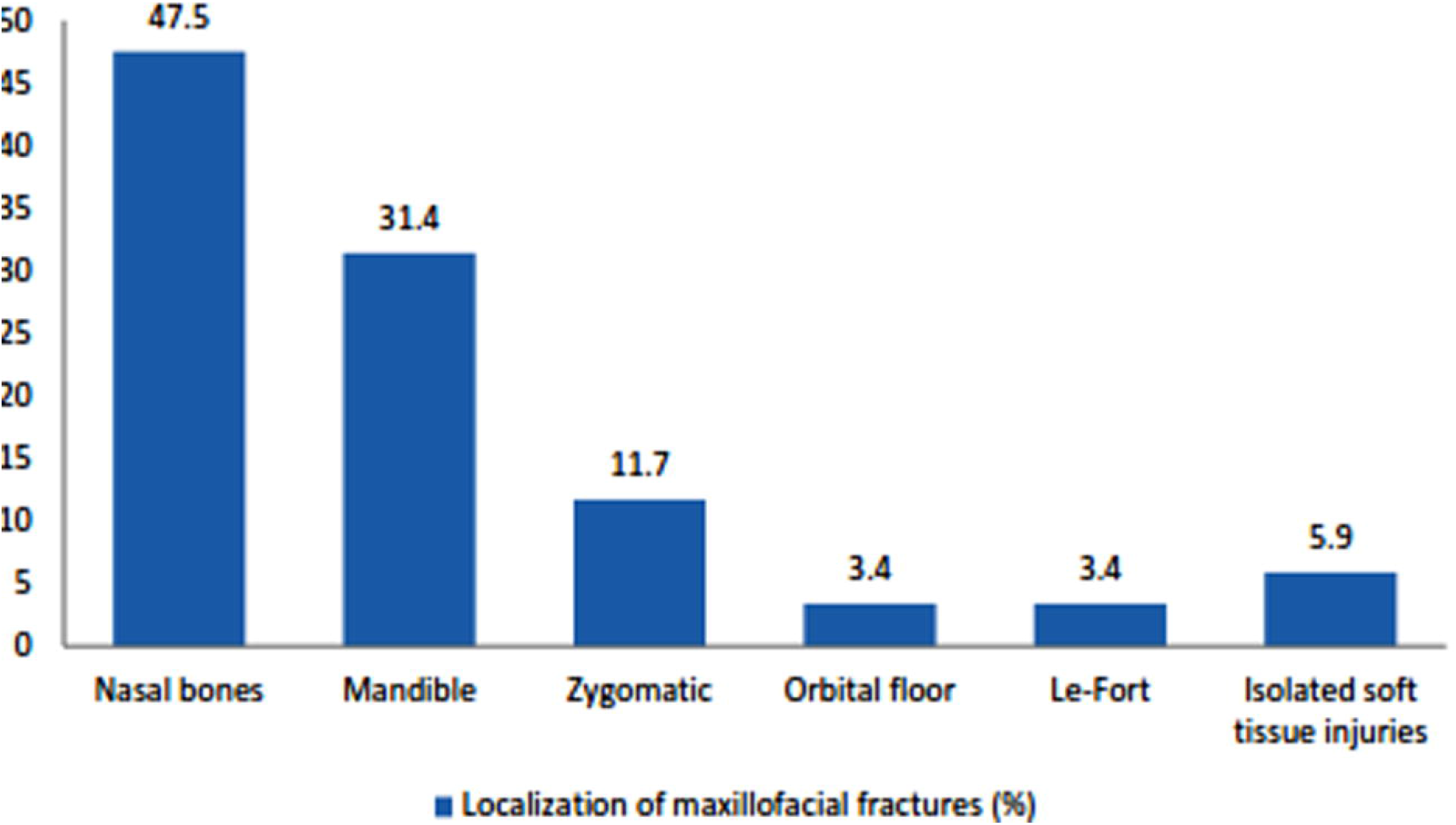

Mandible fractures accounted for 31.4% (64/204) of all MFIs, and the majority of them (71.9%; 46/64) were caused by interpersonal violence (P=0.011, n=64, χ^2^ test). A sex comparison showed a significant higher prevalence of mandible fractures in males (88.9% vs 11.1%; P=0.0052, n=64, χ^2^ test). Bilateral fractures of the mandible were observed in 60.9% of the patients (39/64), and unilateral fractures were observed in 37.5% of the patients (24/64). The total number of mandible fracture sites was 103. The most frequent injury location of mandible fractures was the angle (37.9%), followed by the symphysis/parasymphysis (28.1%) and the body (12.6%). Condyle fractures accounted for only 10.7% of mandible fractures (Figure 5).

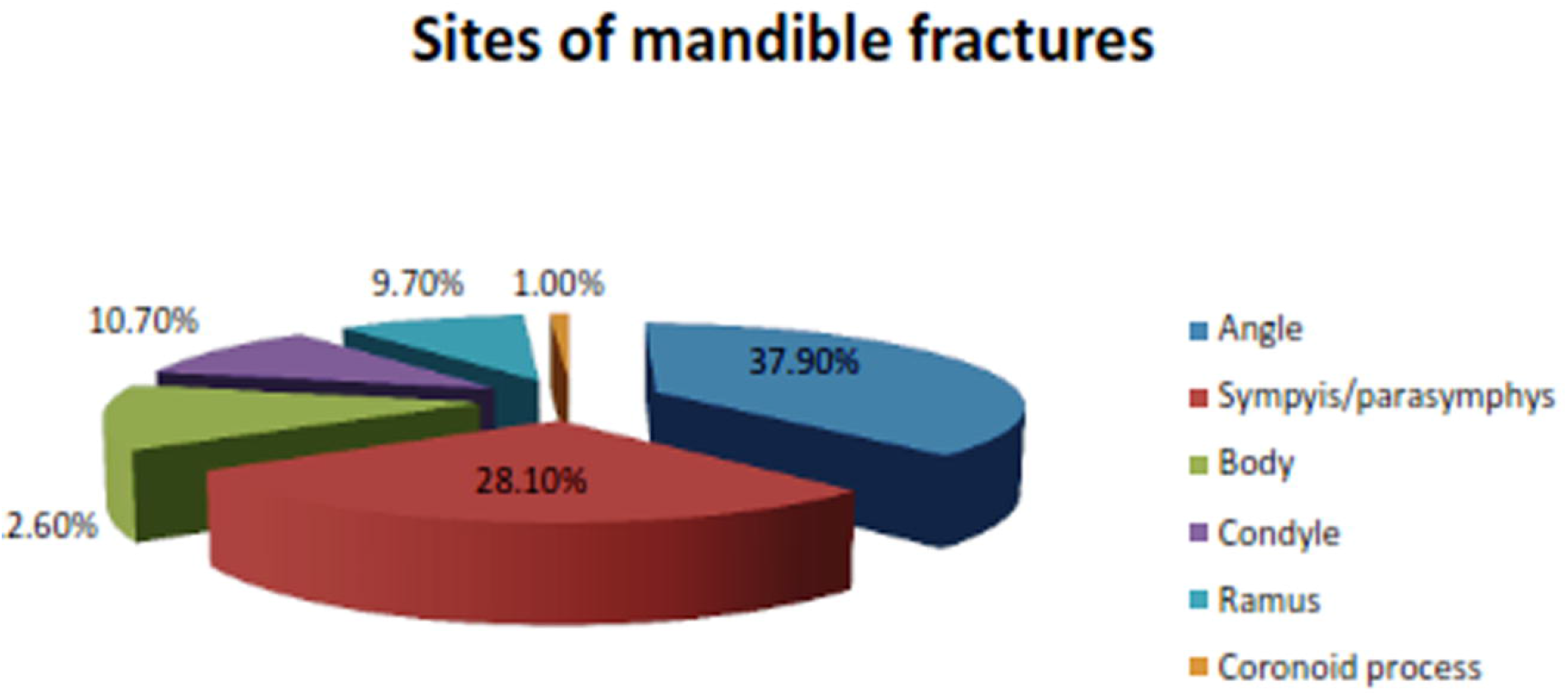

Zygomatic fractures (zygomatico-orbital, zygomatico-maxillary, zygomatico-ethmoidal) accounted for 11.7% (24/204).

Le-Fort fractures were reported in seven (3.4%) cases, of which six were due to RTAs and one was due to an industrial injury. Orbital floor fractures accounted for 3.4% (7/204) of the total number of injuries. Isolated soft tissue injuries were reported in 5.9% (12/204) of the cases (Figure 4).

Combined craniomaxillofacial trauma was observed in 7.8% of the injuries (16/204).

The 2.94% of the fractures (6/204) were treated conservatively. Close reduction accounted for 51.9% of fracture treatments (106/204), 96 of which comprised repositioning of the nasal bones and 10 of which comprised close reduction of the zygomatico-maxillary fracture and zygomatic arch. A total of 42.6% of the fractures (87/204) were treated by ORIF, of which 62 were mandible fractures and 25 were mid-face fractures (zygomatic and Le-Fort fractures) (Figure 6).

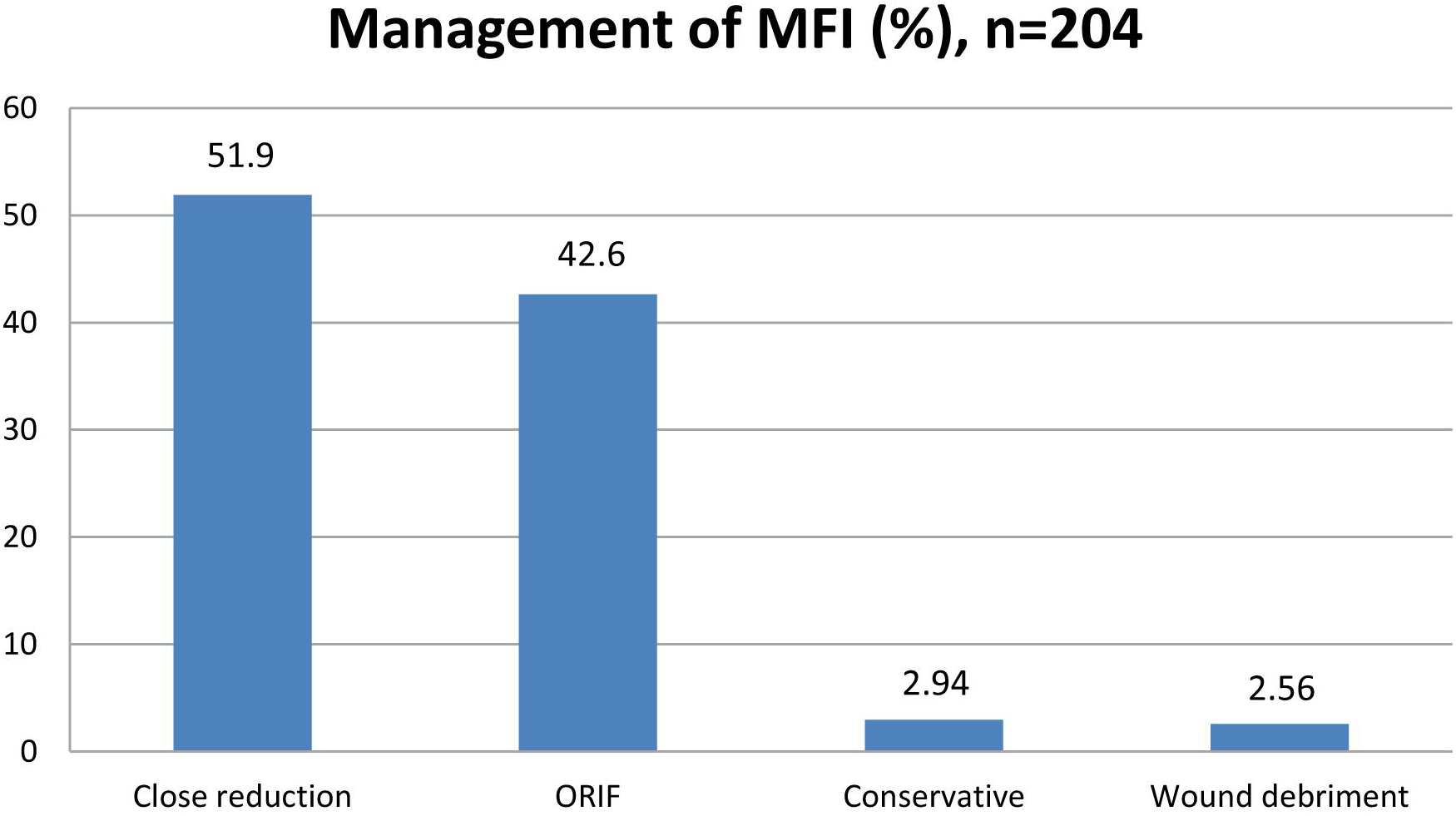

## DISCUSSION

As per the World Bank’s classification, Armenia is a developing country with an upper middle-income economy. The population of Armenia was 2,296,243 in 2020.

Maxillofacial fractures (MFFs) not only cause serious physiological injuries but also impose serious burdens on society due to morbidity, mortality, facial disfigurement, loss of function, and financial expenditures associated with such injuries.[4, 16-18] The incidence rates, etiologies, types, and injuries associated with MFFs vary among different countries and even different areas within the same country due to environmental, socioeconomic, cultural, and lifestyle differences among people.[4, 7, 16]

The proportion of males affected by MFFs in this study was higher than that of females, at 3.25:1, which is in agreement with findings reported in most other studies.[1, 2, 4, 9, 19-22]

IV was found to be the most common etiology of MFF in this study (42.1%, n=190), followed by RTAs (27.9%, n=190) and falls (18.6%, n=190). Most studies on the etiology of maxillofacial trauma in developing countries indicate that RTAs are the most frequent cause of MFIs.[3, 5, 6, 10, 17] In contrast, the most frequent cause of MFFs in developed countries is IV or assault.[1, 6, 10, 15, 23-25]

Sbordone et al.,[26] in their multicentric retrospective study in southern Italy in 2018, showed that the most frequent cause of facial injuries was assault (30.4%), followed by RTAs (27.2%) and falls (23.2%). Boffano et al. [1] analyzed the demographics, causes and characteristics of maxillofacial fractures managed in several European oral and maxillofacial surgery departments over one year. The data of 3396 patients (2655 males and 741 females) with 4155 fractures were recorded and revealed that the most frequent cause of injury was assault, accounting for injuries in 1309 patients; assaults and falls alternated as the most important etiological factors at various centers. The results of the EURMAT collaboration confirmed the changing trend in maxillofacial trauma epidemiology in Europe, with trauma cases caused by assaults and falls outnumbering those due to RTAs.[1] Similar results were observed by Afrooz et al. [24] in their study on the epidemiology of mandibular fractures in the United States. They found that the mechanism of injury differed by sex, with men most frequently sustaining mandibular fractures from assaults (49.1%), followed by motor vehicle accidents (MVAs; 25.4%) and falls (12.8%); women most frequently sustained mandibular fracture from MVAs (53.7%), followed by assaults (14.5%) and falls (23.7%). Falls were a significantly more common etiology in patients who were 65 years or older. Therefore, the MFF epidemiology data obtained in the present study are comparable with data from Europe and the United States.

The 21-40 years age group had the highest MFI incidence rate in the present study. These data are in accordance with data obtained by many other researchers.[6, 7, 9, 15, 17, 19, 23, 27] The main etiological cause of injuries in the 21-30 years age group was IV, followed by RTAs. The high rate in this age group may be due to participation in outdoor activities or psychosocial problems that may provoke risk-taking behaviors, thus making this population more prone to injuries.[28] In this study, patient age was found to be associated with the fracture site. It was demonstrated that patients aged 21-30 years were likely to have sustained nasal bone fractures (40.5%) and mandible fractures (40.5%) in equal proportions. The lowest MFI rate was observed in the elderly age group (>60), with the main etiology of injuries in this group being falls (65%, 13/20).

The most common MFF site and type following trauma varied among studies. The results from most studies showed that the mandible was most commonly affected area.[6, 7, 9, 15, 17, 20, 21, 26, 27] However, in this study, the nasal bones were found to be the most common injury site (47.5%, n=204), followed by the mandible (31.4%, n=204) and zygomatic complex (11.7%, n=204). Comparable data presented by Rezaei et al. [27] in a retrospective study of epidemiology of maxillofacial trauma in a university hospital in Kermanshah, Iran, observed nasal fracture to be the most frequent type of trauma (45.5%), followed by mandibular (29%) and zygomatic (24.9) fractures. The dominance of nasal bone injuries compared to other sites was also noted by Agnihotri et al. [29] who found that the most common bone to be affected was the nasal bone (23.7%), followed by the mandible (22.7%) and zygoma (19.3%). However, the percentage of nasal fractures was two times less than that in the present study, at 23.7% in their study and 47.5% in the current study. The zygoma was the most fractured anatomical site in both males and females in the study by Arangio et al. [20], accounting for 32% of all injuries, followed by isolated fracture of the orbital floor, at 11%. Singaram et al. [10] conducted a retrospective study and showed that 41.9% of fractures were zygoma and maxillary bone fractures, 33.0% were mandibular fractures, 26.2% were dentoalveolar fractures, 8.6% were orbital floor fractures and only 6.4% were nasal bone fractures.

Mandible fractures ranked second among all MFIs in the present study (31.4%, n=204). The most common fracture site was the mandibular angle (37.9%, n=103), followed by the symphysis/parasymphysis (28.1%, n=103) and body (12.6%, n=103). A similar finding on mandible fracture loci distribution was presented by Morris et al.,[19] with the angle accounting for 27%, symphysis accounting for 21.3%, and condyle and subcondyle accounting for 18.4%. Additionally, Ferrer et al. [23] found the most common fracture site to be the mandibular angle (35%), followed by the parasymphysis (30%). Afrooz et al. [24], Kaura et al. [9] and Abhinav et al. [15] noted that the most common site of mandible fracture was the parasymphysis. The results of the EURMAT collaboration by Boffano et al. [1] revealed condylar fracture as the most commonly observed type of mandibular fracture, accounting for 34%, followed by body fractures, angle fractures and fractures of the symphyseal region.

Combined mandible fractures accounted for 59.4% (38/64), and the most frequent association in the present study was the angle and the parasymphysis.

MFFs can be treated with either closed reduction (conservative) or ORIF (surgical) methods or a combined approach. The decision regarding treatment depends on a variety of factors, such as the nature of the injury, the presence of associated injuries and comorbidities, the skill of the surgeon, etc. In the present study, close reduction was performed in all patients with nasal bone fractures and ten patients with minimally displaced zygomatico-maxillary and zygomatic arch fractures. A total of 42.6% of the fractures were treated by ORIF.

## CONCLUSION

Interpersonal violence, followed by RTAs and falls, was the most common cause of MFIs. The nasal bone was the most common injury site, followed by the mandible and zygomatic complex. Social education with the objective of reducing aggression and interpersonal conflict should be improved, and appropriate RTA prevention strategies should be strengthened and implemented.

## Data Availability

All data relevant to the study are included in the article or uploaded as supplementary information

## Data Availability

All data relevant to the study are included in the article or uploaded as supplementary information

## Data Availability

All data relevant to the study are included in the article or uploaded as supplementary information

## ACKNOWLEDGEMENTS

None.

## AUHTOR CONTRIBUTIONS

Poghosyan A.Yu., Tadevosyan A.E., Aleksanyan L.V., Misakyan M.S, Minasyan A.M., Bablumyan A.Yu., Muradyan A.A. makes substantial contributions to conception and design, and acquisition of data, and analysis and interpretation of data; Poghosyan A. Yu., Minasyan A.M., Bablumyan A.Yu., Misakyan M.S., Muradyan A.A., participates in drafting the article or revising it critically for important intellectual content; Tadevosyan A.E., Aleksanyan L.V. performed data curation, methodology; Poghosyan A. Yu., Minasyan A.M., Bablumyan A.Yu., Muradyan A.A., give final approval of the version to be submitted and revised version.

All authors read and approved the final manuscript

## PATIENT CONSENT

Not required for this type of study

## DATA AVAILABILITY

The data that support the findings of this study are available from the corresponding author, upon request.

